# The Impact of Coronavirus Disease 2019 (COVID-19) on Liver Injury in China: A Systematic Review and Meta-analysis

**DOI:** 10.1101/2020.05.03.20089557

**Authors:** Xin Zhao, Zehua Lei, Fengwei Gao, Qingyun Xie, Kangyi Jang, Jianping Wu, Jinqiang Fu, Bo Du, Zhixu Wang

## Abstract

**Background:** The evidence for the incidence and severity of liver injury in Chinese patients with COVID-19 is still controversial.

**Aims:** The purpose of this study was to summarize the incidence of liver injury and the differences between liver injury markers among different patients with COVID-19 in China.

**Methods:** Computer searches of PubMed, Embase, CNKI and medRxiv were used to obtain reports on the incidence and markers of liver injury in Chinese patients with COVID-19, from January 1, 2020 to April 10, 2020. (No. CRD42020181350)

**Results:** A total of 57 reports from China were included, including 9889 confirmed cases of COVID-19 infection. The results of the meta-analysis showed that among the patients with early COVID-19 infection in China, the incidence of liver injury events was 24.7% (95% CI, 23.4%-26.4%). Liver injury in severe patients was more common than that in non-severe patients, with a risk ratio of 2.07 (95% CI, 1.77 to 2.43). Quantitative analysis showed that the severe the coronavirus infection, the higher the level of AST, ALT, TB, ALP, GGT and the lower the level of ALB. The changing trend of the appeal index was similar in ICU patients and dead patients.

**Conclusion:** There is a certain risk of liver injury in Chinese patients with COVID-19, and the risk and degree of liver injury are related to the severity of COVID-19.

## Introduction

In December 2019, an unexplained viral pneumonia broke out in Wuhan, China, and the World Health Organization named it COVID-19[1]. As of April 20, 2020, the cumulative incidence of global COVID-19 has exceeded 2 million, and the number of death is close to 200000[1–3]. Studies have shown that the pathogen of COVID-19 is β-coronavirus, and its gene sequence is highly similar to that of SARS and MERS[4]. The source and specific route of transmission of the virus are still unclear. The principal target organ of COVID-19 is the human lung, and some studies have shown that the virus can damage the heart, liver, nervous system, and so on[5–7]. Liver injury was also noted in SARS and MERS. Guan et al reported that abnormal elevation of AST accounted for 22.19% of COVID-19 patients (168/757). ALT accounted for 21.32% (158/741). Among the 99 cases reported by Chen et al, there were 43 cases of abnormal liver function. These findings seemed to imply that there was a definite relationship between novel coronavirus and liver injury [8–10]. However, the effect of COVID-19 on liver injury is still unclear and requires further study.

AST, ALT, TB, and ALB are important markers for the evaluation of liver injury. There are significant differences in the proportion and degree of increase in AST and ALT in the reports of early COVID-19 patients. China, at the outbreak point of the epidemic, published a large number of research reports in the early stage of the outbreak, including information about COVID-19 liver injury [9,10]. At present, there is no systematic review or meta-analysis of the effects of COVID-19 on liver injury, so the purpose of this study is to perform a meta-analysis to systematically review and analyse the effects of COVID-19 on liver injury in China to provide some reference for clinical practice..

## Methods

The systematic review and meta-analysis has been registered on Prospero (Registration number CRD42020181350; https://www.crd.york.ac.uk/PROSPERO). We reported this systematic review and meta-analysis according to MOOSE guidelines and PRISMA [11, 12].

### Search strategy

Three electronic databases, PubMed, Embase, CNKI and the medRxiv system (https://www.medrxiv.org), were searched by our team. The keywords included “coronavirus”, “nCoV”, “SARS-CoV-2”, “COVID”, “COVID-19”, “NCP” and “China”. We screened the retrieved literature, and the eligible study was to report the occurrence of liver injury or the abnormal rise or abnormal changes of biochemical markers of liver injury (AST, ALT, TB and so on), and the original data should come from China.

### Studies selection

We selected studies that met the following inclusion criteria: (1) nucleic acid-confirmed and clinically confirmed COVID-19 patients; (2) reported studies of liver injury events or liver injury markers; (3) the language was confined to English or Chinese; (4) the source of the research was limited to China; (5) the retrieval time was limited from January 1, 2020 to April 10, 2020. If the research was from multiple centres, we tried to divide it into a single centre for analysis. If there were multiple studies on the same team, the latest research from the team was used for analysis. The following studies were excluded: (1) COVID-19 patients without nucleic acid diagnosis or clinical diagnosis; (2) reports of special groups such as pregnant women and children; (3) studies that only report deaths or critically ill patients; (4) studies that do not report liver injury events or markers of liver injury; (5) research reports that the participants are not from China.

### Data extraction and quality assement

Three authors (ZX, LZH, GFW) performed a preliminary screening of the search literature, and finally (ZX, LZH) two authors read the full text and agreed on which studies to include in the final analysis. The four authors (ZX, GFW, XQY, JKY) independently extracted the relevant information included in the literature, including the first author, the time of publication, the source of the literature, the time of research, the number, age, male-to-female ratio, case distribution, complications, use of therapeutic drugs, events of liver injury, changes in markers of liver injury, and differences in the process of data extraction. Disagreements were settled by discussion of all the investigators. We used the Newcastle-Ottawa Quality Assessment Scale for retrospective studies to assess the methodological quality of included studies.

### Statistical analysis

We used stata16.0 for data analysis. We define liver injury as AST or ALT or TB above the upper limit, as if there are three or two items of data in a study. The term with the highest number of events was used as the analytical data. If the P-value of liver injury is 0.3 to 0.7, meta-analysis was carried out directly; on the contrary, the data were converted by Freeman-Tukey double arcsine transformation to make them in accordance with a normal distribution and then analysed and combined[13]. The ultimate result after data conversion was restored by using the formula P = [sin (tp/2)]^2^. The degree of heterogeneity was evaluated by the Q^2^ and I^2^ indexes. I^2^ is low heterogeneity when it is 0% to 25%, moderate heterogeneity when it is 26% to 75%, and high heterogeneity when it is 76% to 100%. The fixed effect model was used when I^2^ < 25% P > 0.1; if I^2^ ≥ 25% or P ≤ 0.1, the random effect model was used. If only the median and quartile ranges (Q25 and Q75) were reported, then we assumed that the median was equivalent to the average, and the standard deviation (SD) is (Q75-Q25)/2. We also performed a subgroup analysis of the results with heterogeneity, and publication bias was evaluated by Bgger’s test. A bilateral P-value less than or equal to 0.05 was regarded as statistically significant.

## Results

### Eligible studies and Characteristics

4065 articles were obtained by preliminary search, 3754 articles were obtained after repetition, 3478 reading topics and abstracts were excluded, and 274 full-text articles were excluded. After discussion, the three authors (ZX, LZH, GFW) excluded 217 articles, and finally included 57 studies[2, 9, 10, 14–67], all of which were retrospective studies. The detailed search and study selection process are shown in Figure.1. Thirty-five articles were published in public journals, and 22 were published in preprints.

### Study characteristics and quality

Of the 57studies, 23 were from Wuhan and 34 were not from Wuhan. Among them, 5 items were compared between death and survival patients, 4 items were compared between ICU and non-ICU patients, and 39 items were reported between severe patients and non-severe patients, of which the diagnosis of severe and non-severe cases was based on the diagnosis and treatment of pneumonia caused by COVID-19 in China, edition 1-5. A total of 9889 patients diagnosed with COVID-19 were enrolled. The source of the cases was primarily from January to February 2020. Among them, 4606 were male, 4629 were female, and 1618 were critically ill patients. The main comorbidities included hypertension, diabetes, liver disease, and so on. Drug treatment was mainly antibiotics and antivirals. Some studies reported treatment using traditional Chinese medicine, which is shown in Table 1. The Newcastle–Ottawa scale (NOS) was used to evaluate the quality of retrospective studies, the results of NOS score is also shown in Table 1.

### The incidence of liver injury in patients with COVID-19

A total of 33 studies reported liver injury events, and 5867 COVID-19 patients reported liver injury events. The number of liver injuries was 1460, and the initial combined effect was P < 0.3. After converting the data by Freeman-Tukey double arcsine transformation, the meta-analysis obtained a purge of 0.247 [95%CI (0.234, 0.264), P < 0.01], and the incidence of liver injury in Wuhan was 24.7% [95%CI (23.4%-26.4%), P < 0.01] (Supplementary Material, Figure 2). The funnel chart (Supplementary Material, Figure 3) indicated that there was no noticeable deviation in publication. The subgroup analysis of Wuhan and non-Wuhan areas showed that the incidence of liver injury in Wuhan was 26.4% [95%CI (24.3%, 28.2%), P < 0.01] and that outside of Wuhan areas was 23.8% [95%CI (22.1%, 26.0%), P < 0.01]. The incidence of liver injury in Wuhan is slightly higher than that outside of Wuhan(Supplementary Material, Overall Summary of Results Table 2).

### The relationship between the severity of COVID-19 and the incidence of liver injury

Eleven studies reported the occurrence of liver injury caused by increased ALT in severe and non-severe patients, suggesting that the risk of liver injury in severe COVID-19 patients was 1.69 times higher than that in non-severe patients [RR 1.69, 95%CI (1.37, 2.09), P<0.01]. Thirteen studies reported the occurrence of liver injury caused by elevated AST in severe and non-severe patients, suggesting that the risk of liver injury in severe COVID-19 patients was 2.57 times higher than that in non-severe patients [RR 2.57, 95%CI (2.04, 3.25), P<0.01]. Eight studies reported the occurrence of liver injury caused by elevated TB in severe and non-severe patients, suggesting that the risk of liver injury in severe COVID-19 patients was 1.70 times higher than that in non-severe patients [RR 1.70, 95%CI (1.19, 2.44), P<0.01]. The combined analysis showed that the risk of liver injury in severe COVID-19 patients was 2.07 times higher than that in non-severe patients[RR 2.07, 95%CI (1.77, 2.43), P<0.01](Supplementary Material, Figure 4). The funnel chart is symmetrical (Supplementary Material, Figure 5). (Supplementary Material, Overall Summary of Results Table 2).

### The relationship between COVID-19 and the degree of liver injury

In the comparative study of severe and non-severe COVID-19, 27 studies showed that the ALT(U/L) of severe patients was higher than that of non-severe patients[SMD 0.51, 95%CI (0.31, 0.71), p<0.01] (Supplementary Material, Figure 6), There were 29 reports of AST(U/L) comparison. The results showed that the AST of severe patients was higher[SMD 1.05, 95%CI (0.76, 1.34), p<0.01] (Supplementary Material, Figure 7). TB comparison (µmol/L) was performed in 22 items, and the results showed that the TB of severe patients was higher[SMD 0.52, 95%CI(0.34, 0.71), p<0.01] (Supplementary Material, Figure 8). There were seven reports of ALP(U/L) comparisons, and the results showed that the ALP of severe patients was higher[SMD 0.49, 95%CI (0.11, 0.86), p=0.01] (Supplementary Material, Figure 9). There were six reports of GGT(U/L) comparison, and the results showed that the GGT of severe patients was higher[SMD 0.64, 95%CI (0.15, 1.14), p=0.01] (Supplementary Material, Figure 9). The consequences of 20 reports of ALB(g/L) comparison showed that ALB was lower in severe patients[SMD -1.2, 95%CI (−1.47, −0.93), p<0.01] (Supplementary Material, Figure 10). The subgroup analysis of ALT, AST, TB and ALB for severe patients in Wuhan and non-Wuhan areas showed no significant change in heterogeneity (Supplementary Material, Figure 11-Figure 14, Table 2). In 3 reports of ICU and non-ICU and 4 reports of survival and death of COVID-19 patients, the changing trend of ALT, ALT, TB, and ALB was comparable to that of severe and non-severe patients (Supplementary Material, Figure 6-Figure8, Figure 10, Table2). The above results showed that there was no publication bias in the Bgger test, suggesting that the more intense the COVID-19 infection, the more severe the liver injury.

### Publication Bias and Sensitivity Analysis

The funnel plot and Bgger test showed no publication bias for rate of liver injury (Figure 3 and Table 2), the relationship between the severity of COVID-19 and the incidence of liver injury (Figure 5 and Table 2) and the relationship between COVID-19 and the degree of liver injury (Table 2). Forest plot showed little sensitivity change by systematically removing each study for rate of liver injury, the relationship between the severity of COVID-19 and the incidence of liver injury, the relationship between COVID-19 and the degree of liver injury.

## Discussion

In this systematic review and meta-analysis, we found that the incidence of liver injury in Chinese COVID-19 patients was 24.7%, and the incidence of liver injury in Wuhan was slightly higher than that in non-Wuhan patients. At present, the mechanism of liver injury in patients with COVID-19 includes SARS-CoV-2 virus directly attacking liver tissue, and the target receptor of SARS-CoV-2 is the ACE2 receptor. However, previous studies have shown that the ACE2 receptor is highly expressed in intrahepatic bile duct epithelial cells but is expressed at low levels in hepatocytes[68, 69]. Therefore, it is possible for the SARS-CoV-2 virus to directly damage the intrahepatic biliary system, but it is less likely to directly damage hepatocytes. In our study, we also found that GGT is a marker of bile duct injury. ALP increased, but only 7 cases were reported. The mechanism of secondary liver injury includes the inflammatory storm effect caused by systemic inflammation, drug factors and multiple organ dysfunction caused by hypoxia. A meta-analysis by Wang et al[70] concluded that there was no significant correlation between chronic liver disease patients and coronavirus severity. Mantovani et al[71] reported that the total prevalence rate of COVID-19 in patients with chronic liver disease was only 3%. Two other studies reported that liver injury markers in mild patients were at normal levels, which seemed to suggest that COVID-19 was less likely to directly damage the liver[2, 46].

In our study, we observed that the incidence of liver injury and AST, ALT, TB, GGT, and ALP quantitative analysis of liver injury markers in severe patients were higher than those in non-severe patients, while the quantity of ALB in severe patients was lower than that in non-severe patients. The trend of ICU was similar to that of non-ICU patients. This conclusion is consistent with the clinical characteristics of patients with COVID liver injury analysed by Qi et al[7]. All these findings suggest that the more serious the COVID infection, the higher the risk of liver injury and the more severe the injury. The more intense the infection symptoms of critically ill patients are, the greater the impact of the storm of inflammatory factors on liver function. In addition, most of patients included in the study were diagnosed between January and February 2020, who were treated promptly, when the drug treatment of critically ill patients is often uncertain, and antiviral, antibiotic, and traditional Chinese medicine treatments are often combined. It will cause some damage to liver function. Additionally, patients with severe COVID-19 often have hypoxemia, which leads to hepatocyte ischaemia and hypoxia, and multiple organ failure may also be one of the causes of liver damage. Therefore, secondary liver injury is more common in critically ill patients, but the possibility of direct liver injury caused by COVID-19 cannot be ruled out. Therefore, in critically ill patients, real-time monitoring of liver function, evaluation of liver injury, and inexpensive choice of drug treatment are necessary. We also included four analyses of liver damage in dead and surviving COVID-19 patients. Changes in liver injury markers were similar to those of severe and non-severe patients. Owing to data limitations, we did not further analyse whether liver injury increases the risk of death in patients with COVID-19.

Our study also has some limitations. First, although there is no obvious bias in the definition of liver injury and detection of liver injury markers, some of the results are still heterogeneous. Second, this study only analysed the data of Chinese COVID-19 patients, not remote data analysis. Third, it is not possible to further describe the effects of other confounding factors, such as complications, age, and gender, on the results of the study; these confounding factors are also difficult to adjust. Fourth, most of the included studies are retrospective analyses, some are cross-sectional studies, 22 are manuscripts that have not been peer-reviewed, and there is a danger of bias in data collection.

## Conclusion

There is a certain risk of liver injury in Chinese patients with COVID-19, and the risk and degree of liver injury are related to the severity of COVID-19.

## Abbreviations

COVID-19: Coronavirus Disease 2019
ALT: Alanine aminotransferase
AST: Aspertate aminotransferase
TB: Total bilirubin
ALP: Alkaline phosphatase
GGT: γ-glutamyl transpeptidase
ALB: Albumin
CNKI: China National Knowledge Infrastructure

## Data Availability

All data in the manuscript is available

## Conflicts of interest

Nothing to declare.

## Acknowledgements

Thanks for prof. Zehua Lei revising the manuscrip

---

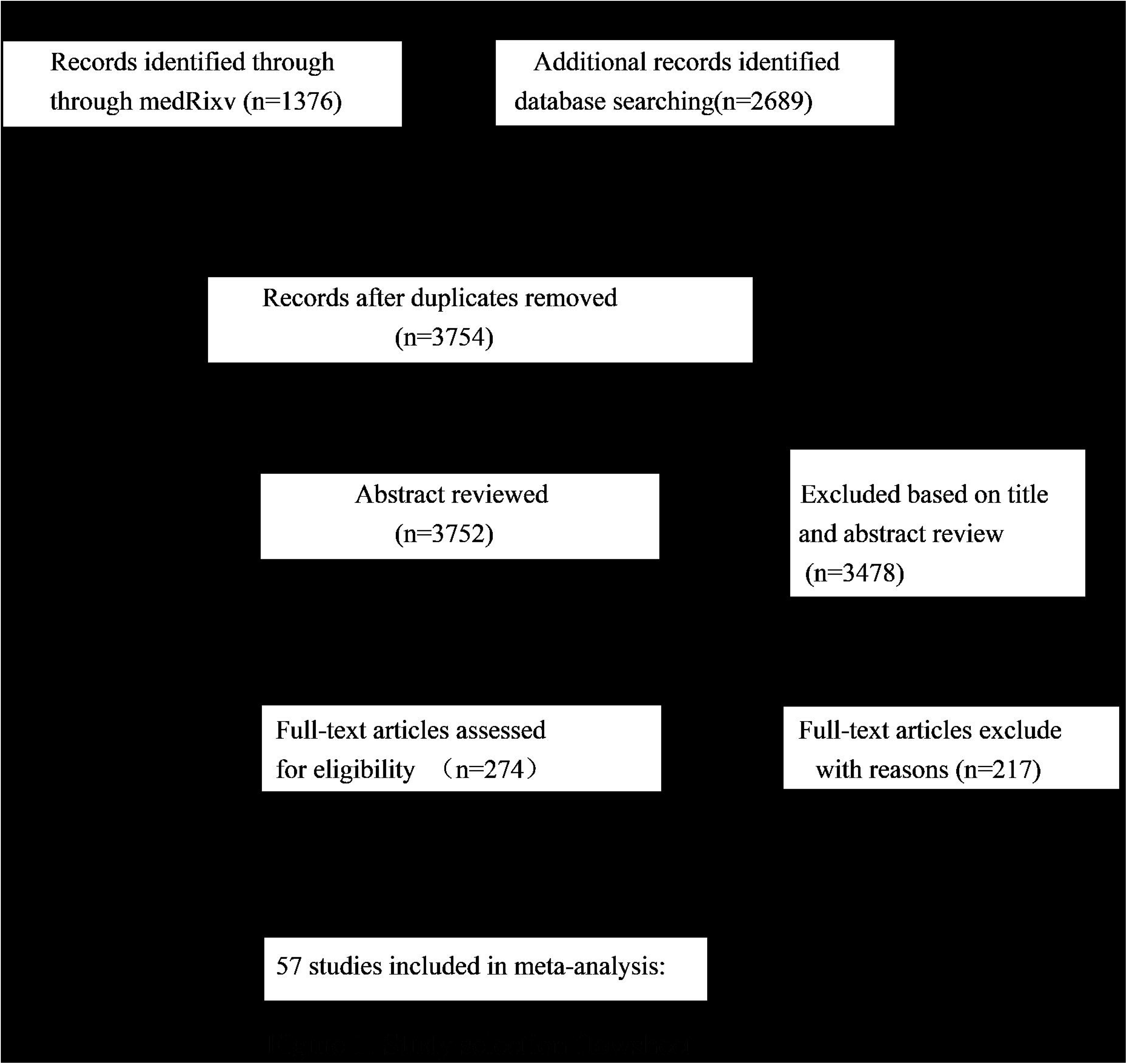

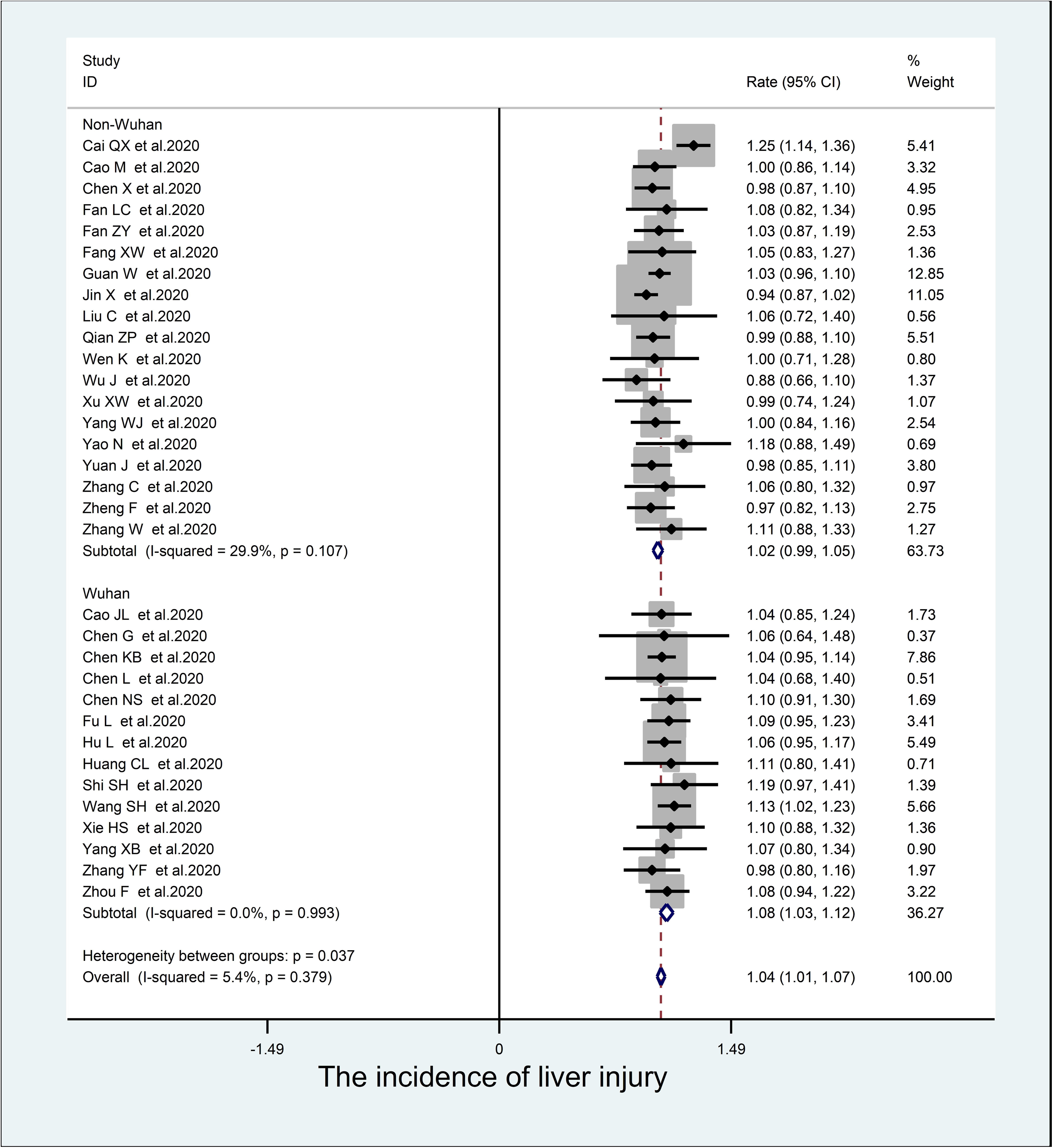

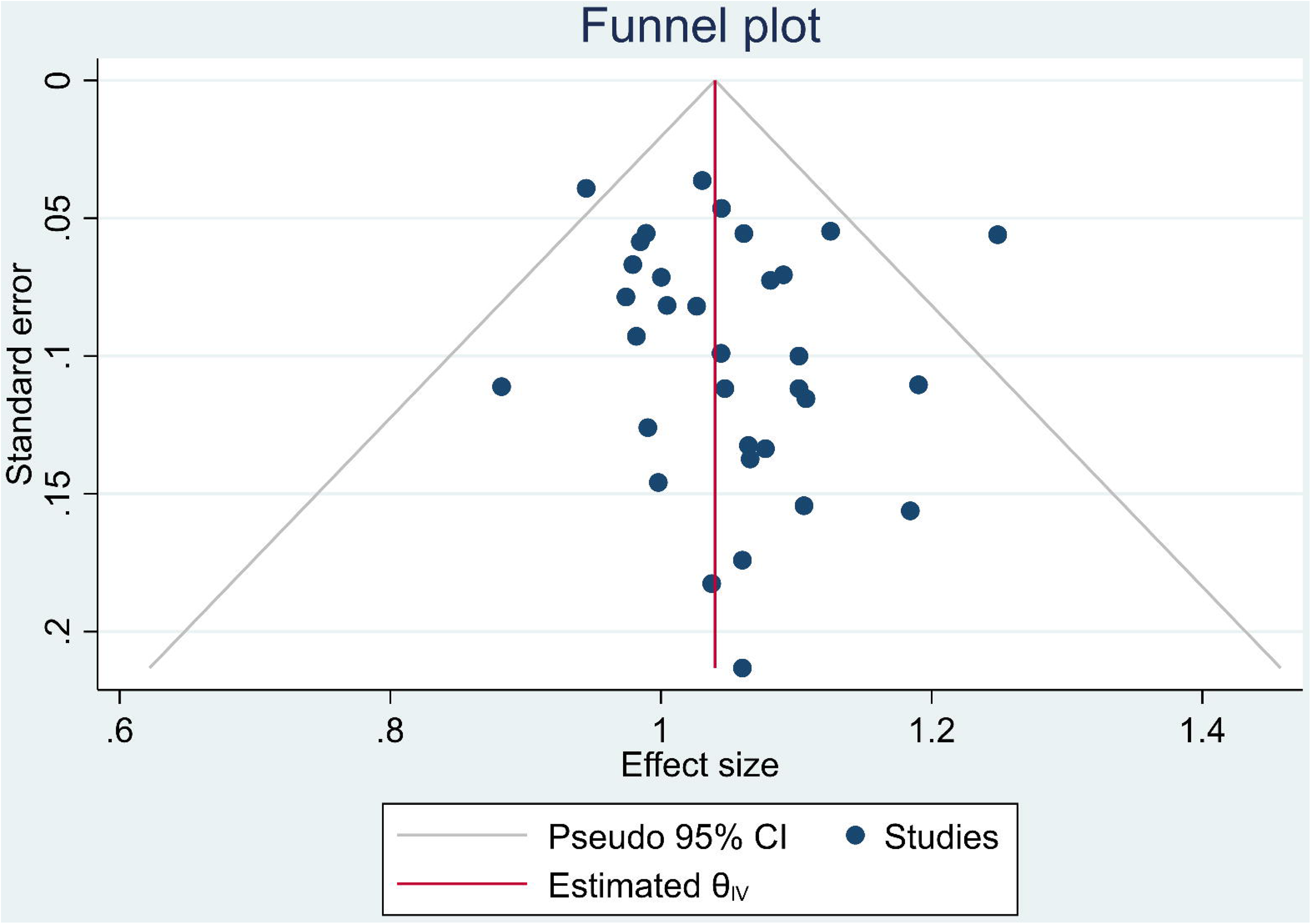

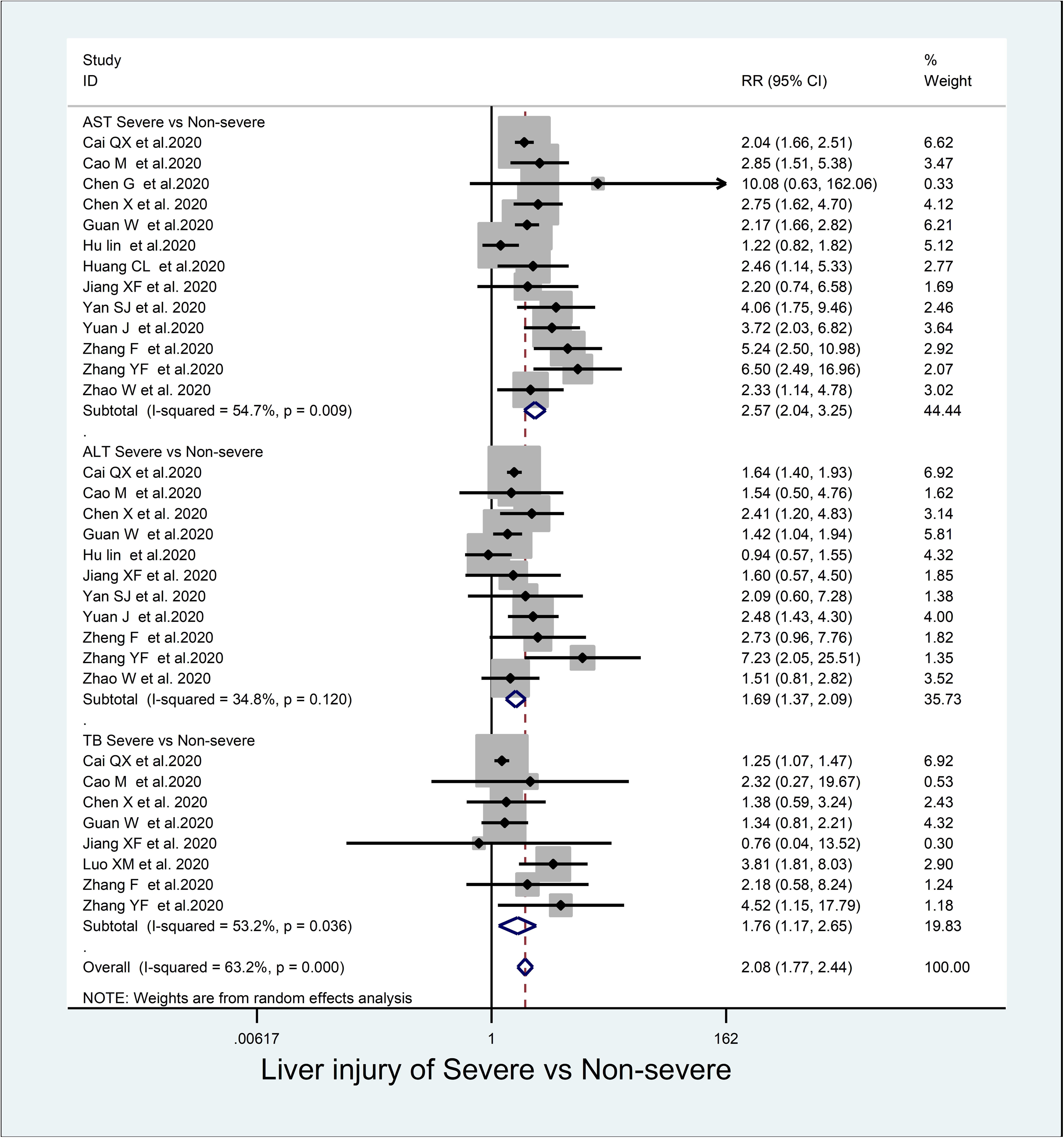

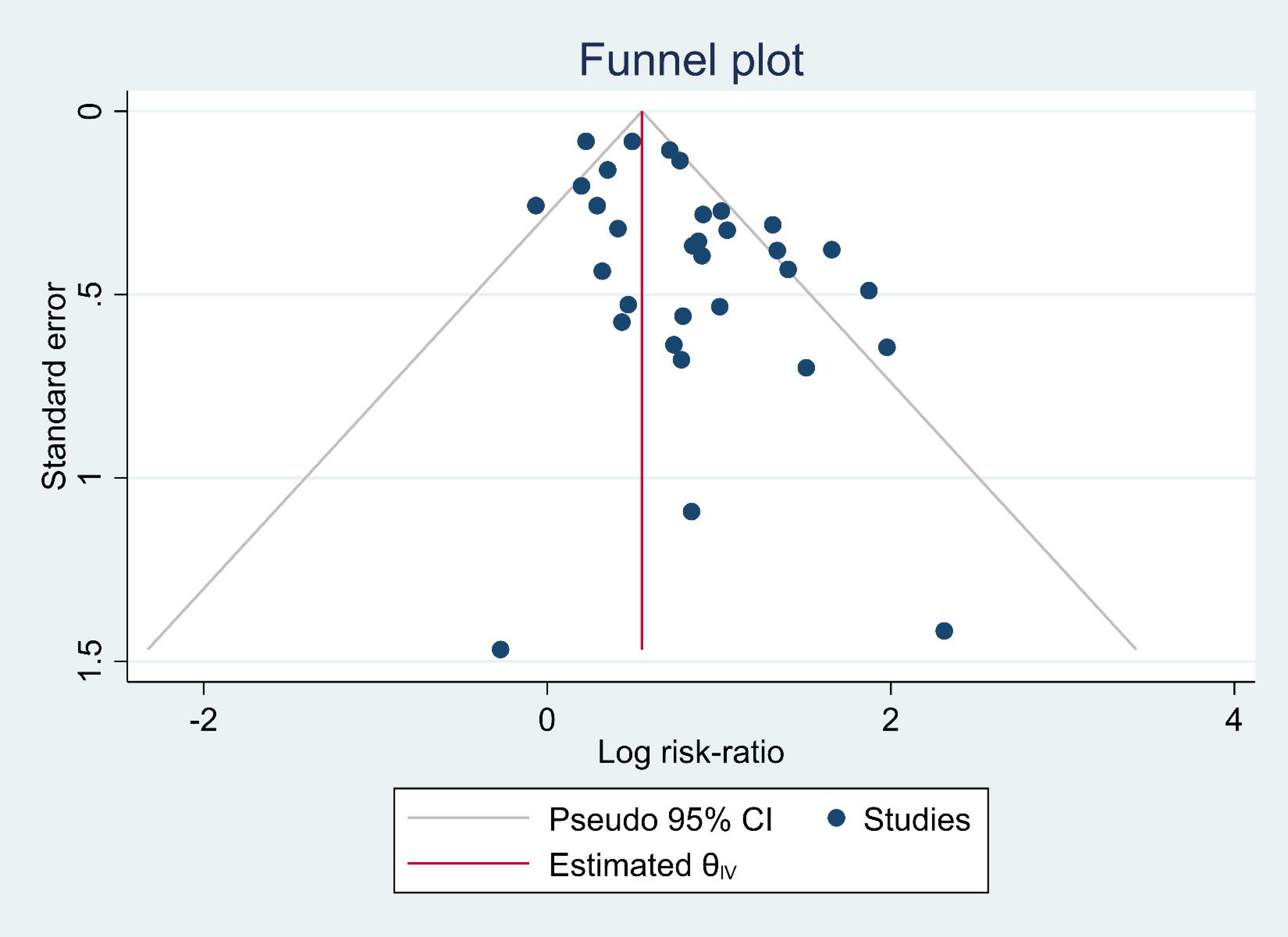

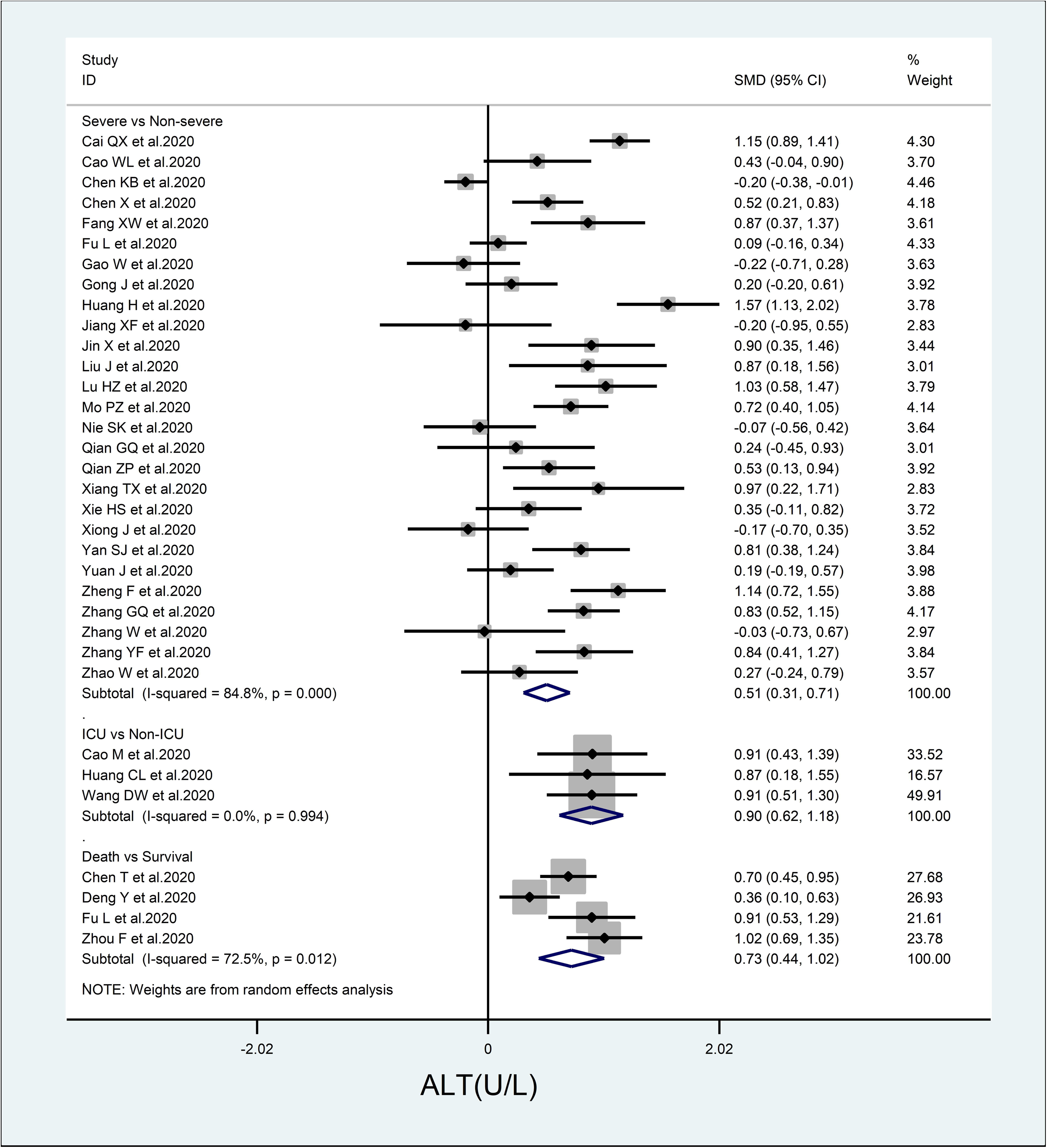

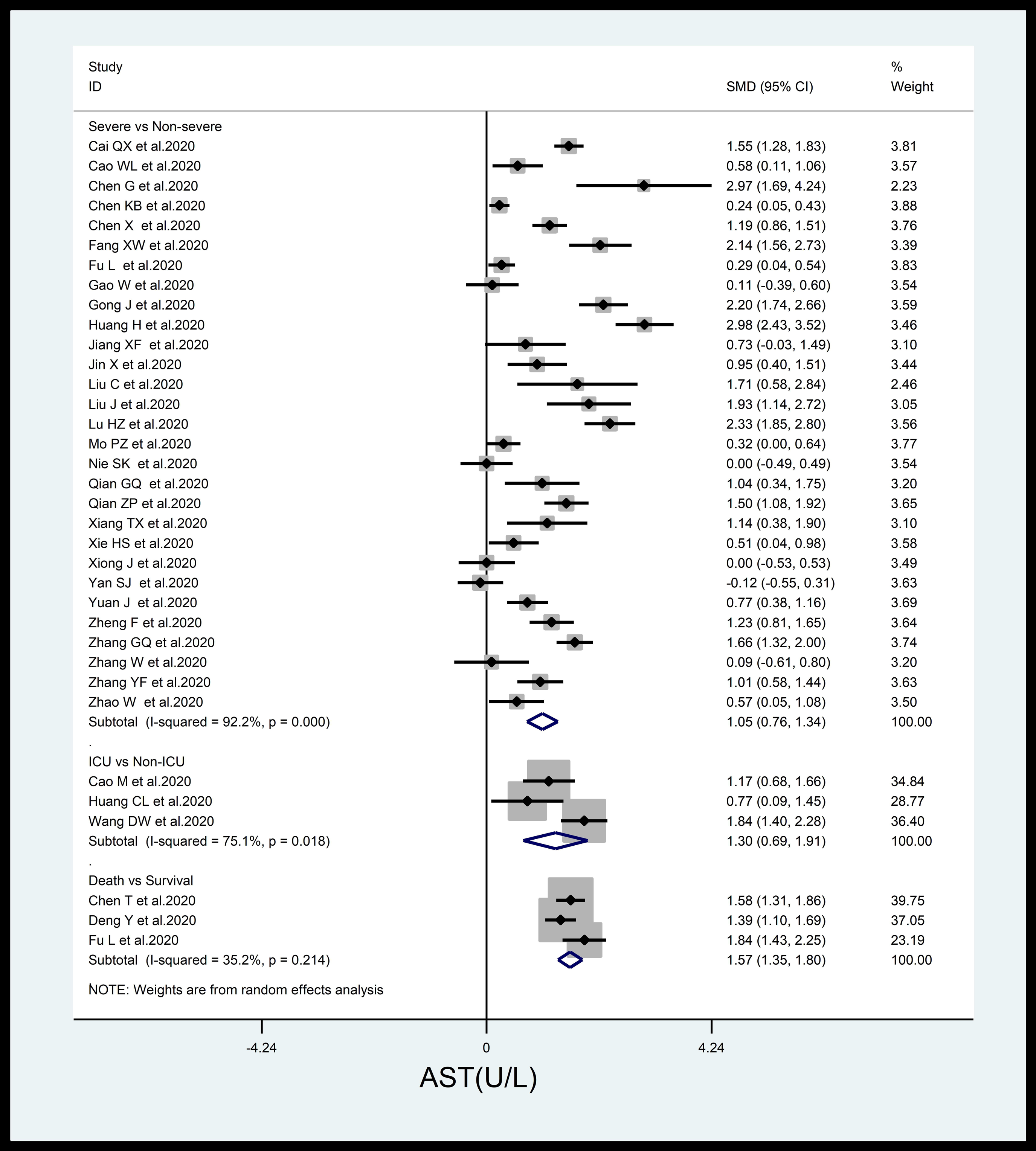

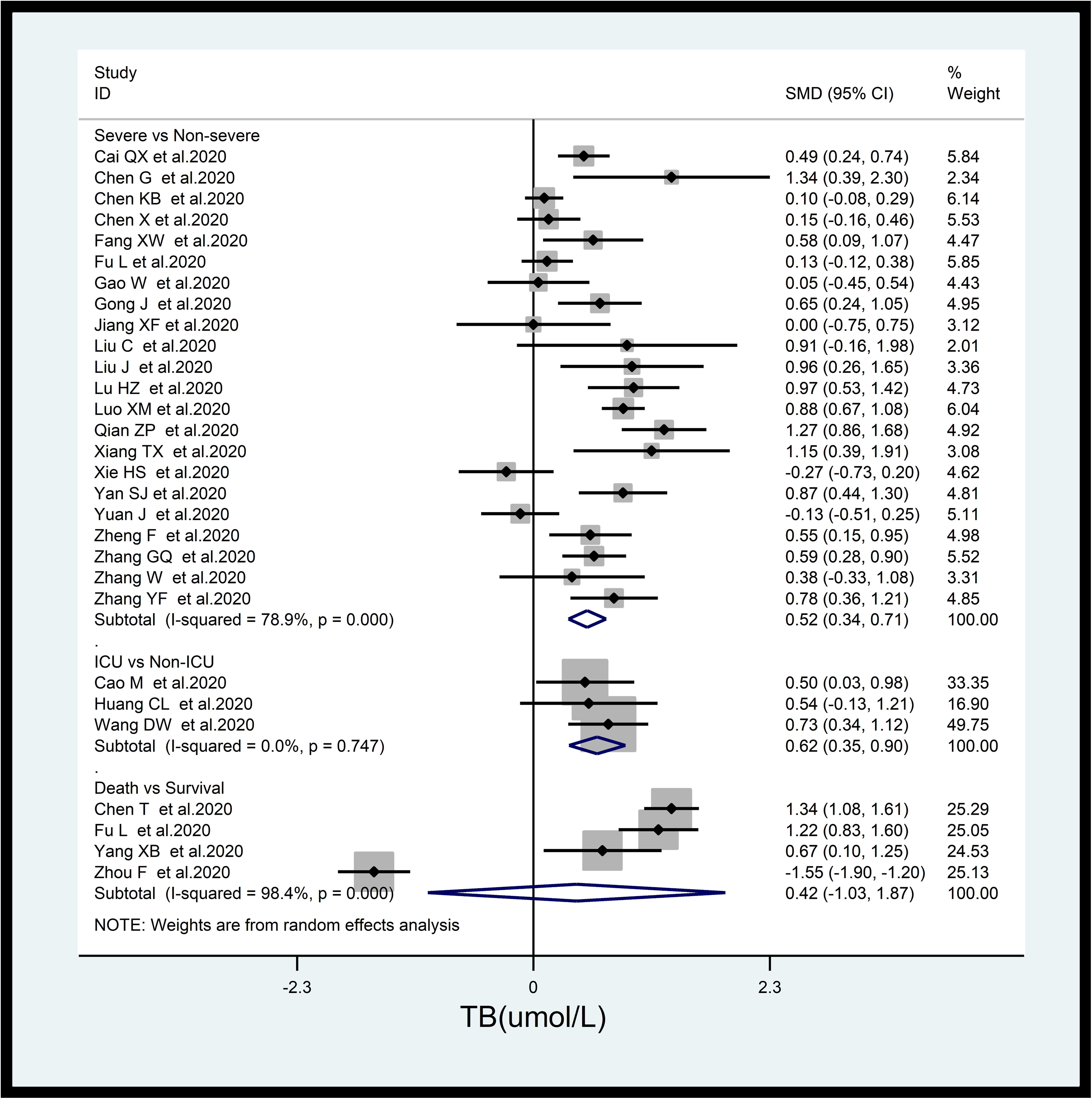

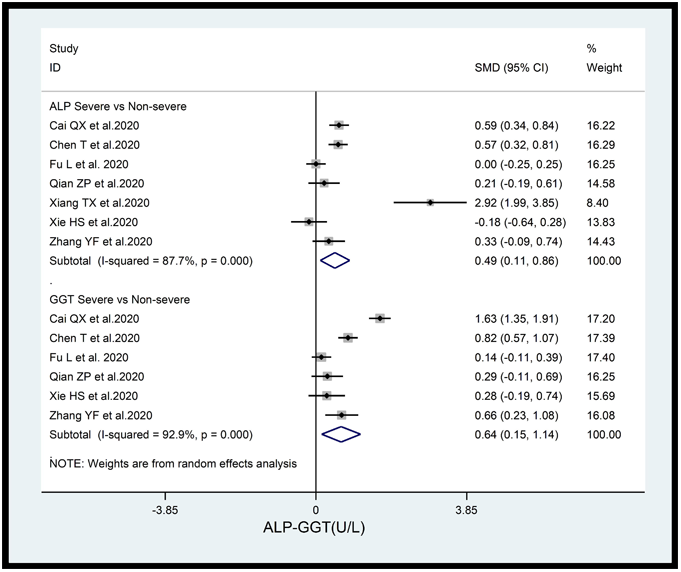

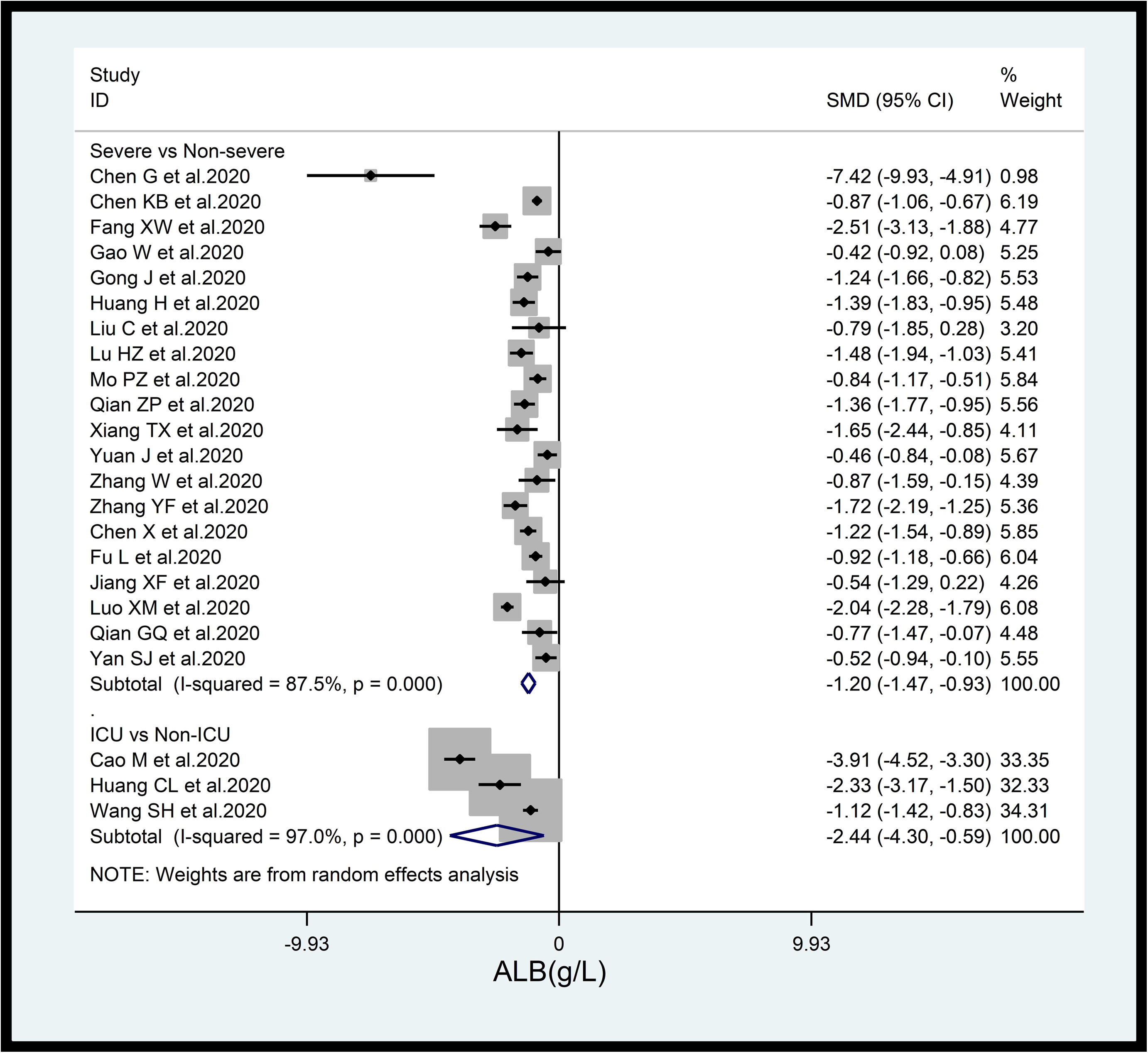

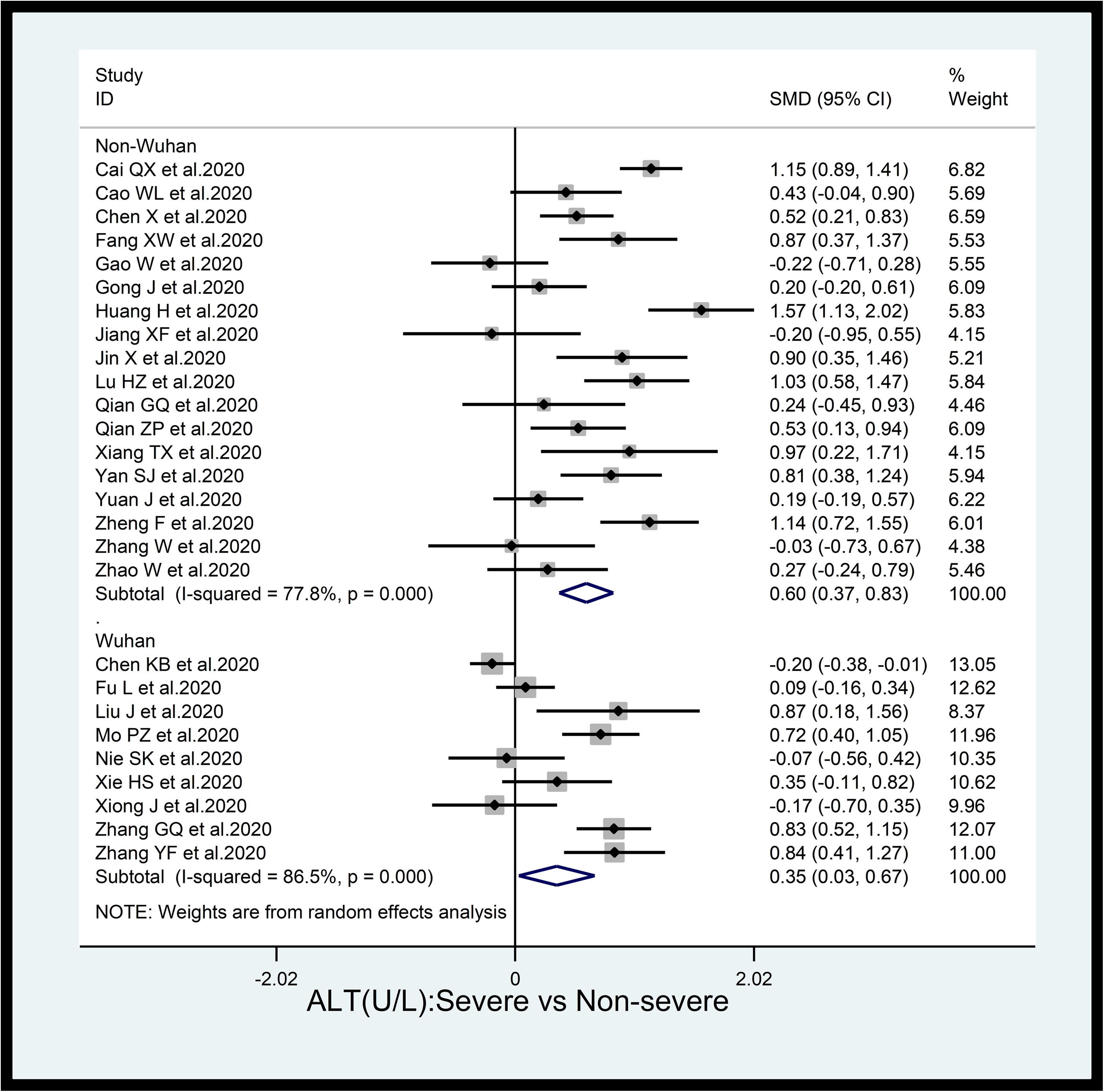

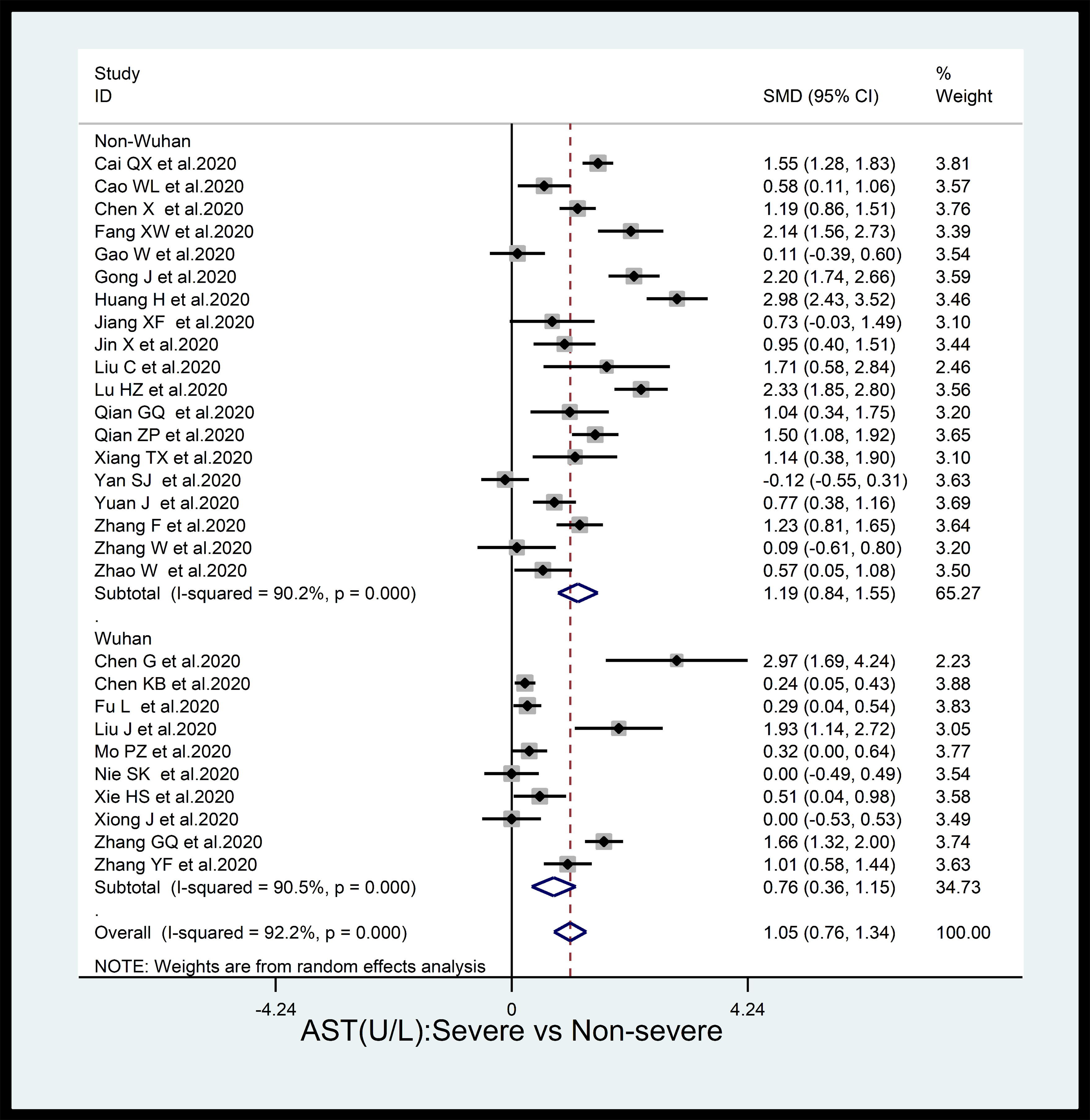

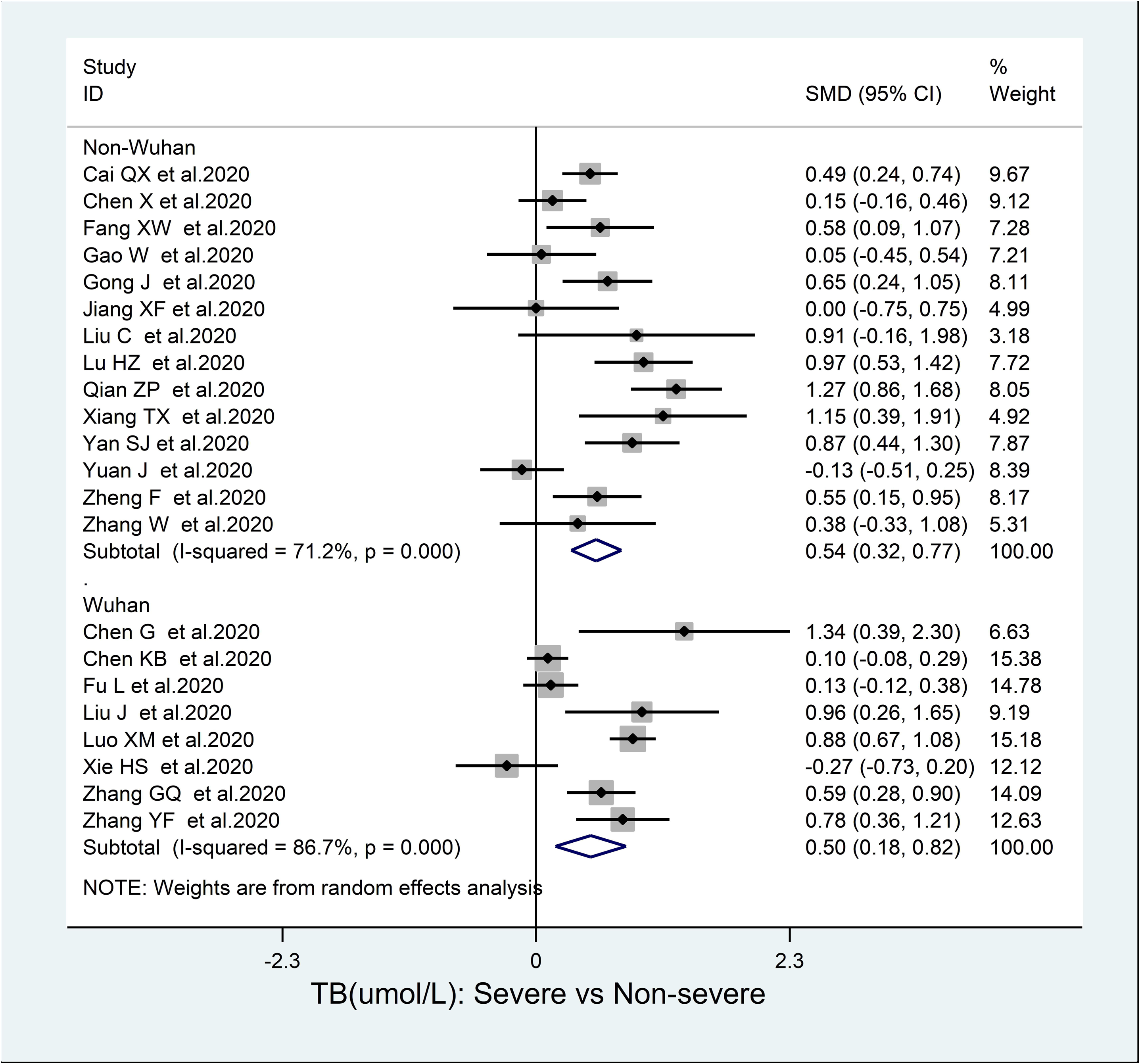

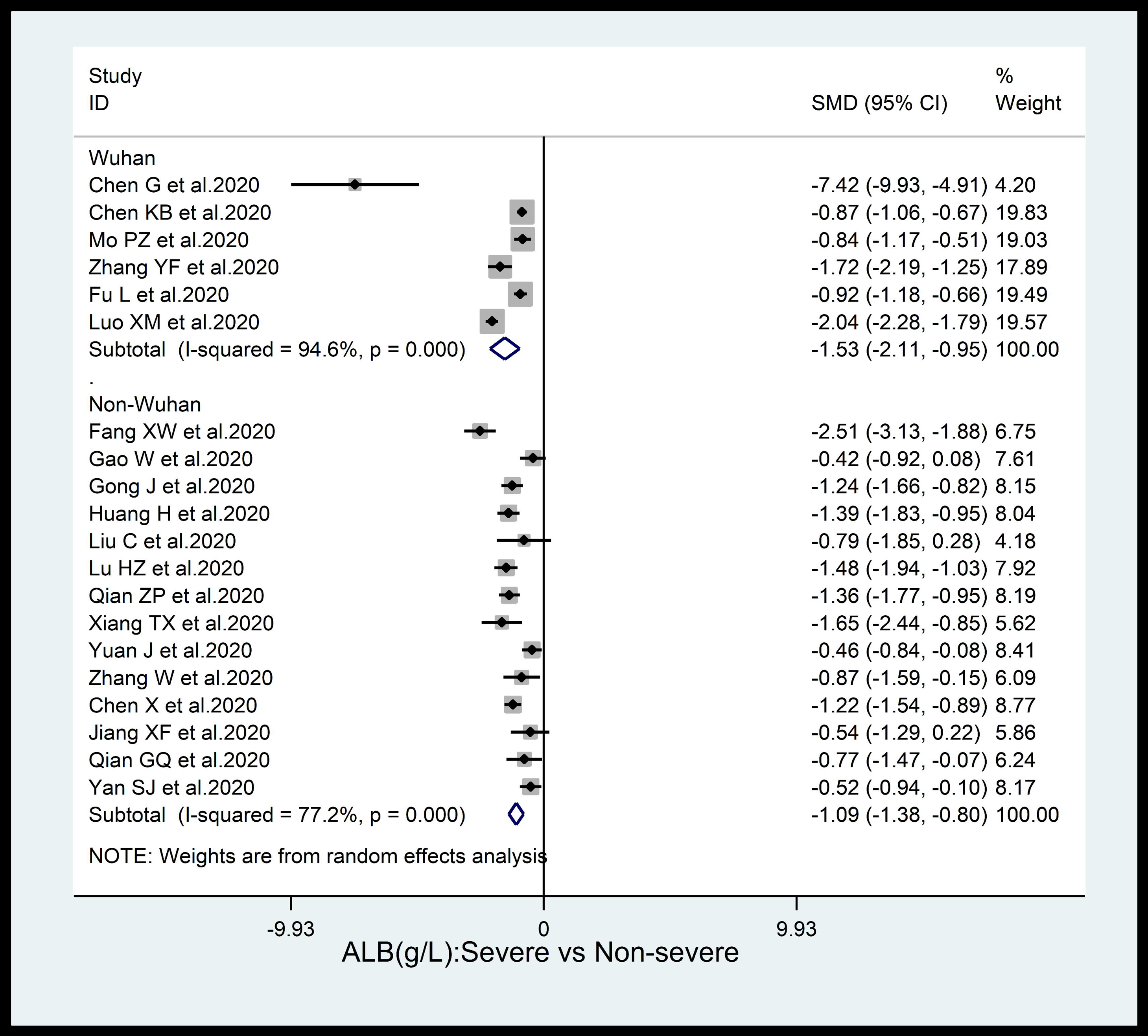

## Notes

**Conflict of Interest Statement** No conflicting relationship exists for any of the authors.

### Competing Interest Statement

The authors have declared no competing interest.

### Clinical Trial

CRD42020181350

### Funding Statement

No funding

